# Mitigating Pathogenesis for Target Discovery and Disease Subtyping

**DOI:** 10.1101/2023.08.12.23294026

**Authors:** Eric V. Strobl, Thomas A. Lasko, Eric R. Gamazon

## Abstract

Treatments ideally mitigate pathogenesis, or the detrimental effects of the root causes of disease. However, existing definitions of treatment effect fail to account for pathogenic mechanism. We therefore introduce the *Treated Root causal Effects* (TRE) metric which measures the ability of a treatment to modify root causal effects. We leverage TREs to automatically identify treatment targets and cluster patients who respond similarly to treatment. The proposed algorithm learns a partially linear causal model to extract the root causal effects of each variable and then estimates TREs for target discovery and down-stream subtyping. We maintain interpretability even without assuming an invertible structural equation model. Experiments across a range of datasets corroborate the generality of the proposed approach.

## I. Introduction

Target discovery refers to the process of identifying disease-modifying targets for the development of novel treatments. Candidate targets should causally affect patient symptoms. We seek to discover treatment targets from data with minimal prior knowledge, time and expense.

Properly identifying treatment targets requires a careful definition of treatment effect. Most investigators quantify treatment effect using counterfactuals or the do-operator found in the causal inference literature [1], [2]. Unfortunately, these quantities ignore the effect of treatment on pathogenesis. Consider for example the following causal graph in a patient with appendicitis:

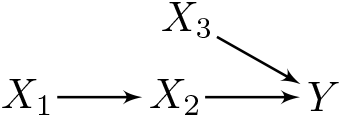

where fecal impaction *X*_***1***_ causes bacterial inflammation of the appendix *X*_***2***_ which in turn causes lower abdominal pain *Y* [3]. We can treat the patient with opioids or other pain medications *X*_***3***_ that directly act on *Y*. However, these medications ignore the pathogenesis of impaction and inflammation leading to the lower abdominal pain. We need definitive treatments that remove the inflammation (e.g., antibiotics) or both the impaction and inflammation (appendectomy). We thus seek a new definition of treatment effect that accounts for the ability of a treatment to modulate pathogenic mechanism.

A pathogenically informed formulation of treatment effect may also assist with diagnosis. Modern clinical diagnoses of complex diseases, such as schizophrenia, fail to map onto the few biological targets needed for potent treatment development [4]–[6]. Investigators have therefore proposed to discover *theratypes*, or disease categories that delineate patients with distinct responses to potentially undiscovered treatments [7]. For example, physicians once categorized anemia as a single disease. Further research revealed the presence of multiple subtypes, such as those responsive to iron and vitamin B12 supplementation [8], [9]. We now categorize iron and vitamin B12 deficiency-induced anemia as two distinct theratypes. This example suggests that we can identify theratypes directly by their differential treatment effects – provided that the treatment effects properly account for pathogenesis.

We make the following contributions in this paper:

1. We summarize the causal effects associated with pathogenesis using the *root causal effects*, or the causal effects of the root causes of disease.
2. We measure treatment effect using the *Treated Root causal Effects* (TRE) metric that quantifies the ability of a treatment to change the root causal effects.
3. We introduce an algorithm that estimates the root causal effects and TREs from observational data under a partially linear model.
4. We employ hierarchical clustering of the estimated TREs to identify theratypes such that grouped patients have pathogenic mechanisms responding similarly to targeted treatments.

Experiments highlight the generality of the approach by demonstrating markedly improved performance across a range of datasets.

## II. Background

We can formally represent a causal process over a set of *p*+1 *endogenous* variables ***X*** using a structural equation model (SEM) linking the variables with deterministic functions and error terms:

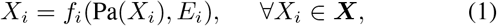

where ***E*** is a set of mutually independent and *exogenous* error terms. The set Pa(*X*_*i*_) ⊆ ***X \*** *X*_*i*_ corresponds to the *parents* of *X*_*i*_. We call *X*_*i*_ a *child* of *X*_*j*_ if *X*_*j*_ ∈ Pa(*X*_*i*_). If Pa(*X*_*i*_) = θ, then *X*_*i*_ is a *root vertex*. We assume *X*_*i*_ = *E*_*i*_ if *X*_*i*_ is a root vertex without loss of generality. We can recover the error term values uniquely from the endogenous variable values in an *invertible* SEM [10].

We associate a directed graph 𝔾 to an SEM by drawing a directed edge from each *X*_*j*_ ∈ Pa(*X*_*i*_) to *X*_*i*_ for every *X*_*i*_ ∈ ***X***. A *directed path* from *X*_*i*_ to *X*_*j*_ corresponds to a sequence of adjacent directed edges from *X*_*i*_ to *X*_*j*_. *X*_*i*_ is an *ancestor* or *cause* of *X*_*j*_, and *X*_*j*_ is a *descendant* of *X*_*i*_, if there exists a directed path from *X*_*i*_ to *X*_*j*_ (or *X*_*i*_ = *X*_*j*_). We collect all ancestors and descendants of *X*_*i*_ into the sets Anc(*X*_*i*_) and Dec(*X*_*i*_), respectively. A *cycle* exists if there is a directed path from *X*_*i*_ to *X*_*j*_, and the directed edge *X*_*j*_ → *X*_*i*_. A directed graph is called a *directed acyclic graph* (DAG) if it contains no cycles. We assume that 𝔾 is a DAG throughout. If we have *X*_*i*_ → *X*_*j*_ ← *X*_*k*_, then we call *X*_*j*_ a *collider*. Two variables *X*_*i*_ and *X*_*j*_ are *d-connected* given ***W*** ⊆ ***X \*** { *X*_*i*_, *X*_*j*_ } in 𝔾 if there exists a path between *X*_*i*_ and *X*_*j*_ such that every collider on the path is an ancestor of ***W*** and no non-collider on the path is in ***W*** . The two vertices are *d-separated* if they are not d-connected. If an SEM associated with a DAG obeys Equation (1), then the joint distribution over ***X*** satisfies the *global Markov property* such that d-separation between *X*_*i*_ and *X*_*j*_ given ***W*** implies conditional independence between *X*_*i*_ and *X*_*j*_ given ***W*** [11].

The *do-operator* do(***A*** = ***a***) represents a *treatment* (also known as an *intervention*), where we manually set the values of ***A*** ⊆ ***X*** to ***a*** by replacing *f*_*i*_ for each *X*_*i*_ ∈ ***A*** in Equation with *X*_*i*_ = *x*_*i*_. We write do(***a***) for shorthand and associate the treatment with the graph 𝔾_do***(a)***_ obtained by removing the directed edges into each member of ***A*** from 𝔾. We have *E*_*i*_ = *X*_*i*_ = *x*_*i*_ for any *X*_*i*_ ∈ ***A*** after the do-operation because ***A*** only contains root vertices in 𝔾_do***(a)***_. The notation do(***A, b***) similarly means that we remove the directed edges into each member of ***A*** ⋃ ***B*** from 𝔾 to create 𝔾_do***(A***,***b)***_. We replace *f*_*i*_ for each *X*_*i*_ ∈ ***A*** in Equation (1) with *X*_*i*_ = *X*_*i*_, and replace *f*_*i*_ for each *X*_*i*_ ∈ ***B*** with *X*_*i*_ = *x*_*i*_.

A linear SEM obeys the following form:

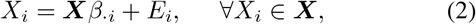

where *β*_*ji*_ ≠ 0 if and only if *X*_*j*_ ∈ Pa(*X*_*i*_). We assume 𝔼 (***X***) = 0. If the error terms follow continuous non-Gaussian distributions, then we more specifically refer to Equation (2) as the Linear Non-Gaussian Acyclic Model (LiNGAM); LiNGAM is invertible [12]. We can rewrite the above equation in matrix form:

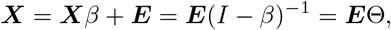

where Θ denotes the matrix of *total effects* of ***E*** on ***X***. Let *θ* refer to the column vector in Θ associated with a target *Y* = *X*_*p****+1***_ ∈ ***X***. We can *augment* 𝔾 by including directed edges from each *E*_*i*_ to *X*_*i*_ except when *X*_*i*_ = *E*_*i*_ is already a root vertex. We display the augmented graph for the appendicitis example below:

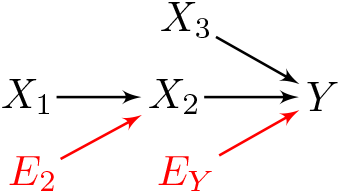

The *root causes* of *Y* correspond to root vertices that are ancestors of *Y* in the augmented graph [13]. If *E*_*i*_ is a root cause of *Y*, then *X*_*i*_ is the projection of the error term onto ***X***, and *E*_*i*_*θ*_*i*_ is the *root causal effect* of *X*_*i*_ on *Y* [14], [15]. We have *X*_***3***_ = 0 (no pain medications) in the appendicitis example even though *θ*_***3***_ = *β*_***3****Y*_ ≠ 0, so that the root causal effect of *X*_***3***_ on *Y* is zero before treatment. Note that we define root causes and their effects relative to the observed variables ***X***; if *X*_*i*_ = *E*_*i*_ is a root cause of *Y* for ***X***, but unobserved *X*_*p****+2***_ has a non-zero total effect on *Y* with *X*_*p****+2***_ → *X*_*i*_, then *X*_*p****+2***_ is a root cause of *Y* for ***X*** ∪ *X*_*p****+2***_ but not for ***X***.

## III. Related Work

We will identify treatment targets by quantifying their ability to modulate root causal effects on a phenotypic response *Y*. However, most investigators currently identify treatment targets with high throughput screening, where they test the causal effects of a large number of molecules on a disease phenotype or target assay [16]. The process incurs substantial cost and time partly because most high throughput screens ignore the pathogenic mechanisms underlying the disease.

Investigators have thus also designed algorithms that identify treatment targets by incorporating knowledge of biological networks [17], [18]. Many methods represent the network using an undirected graph, where edges denote statistical associations or binding affinities. Scientists then utilize measures of proximity or centrality to predict treatment effect [19]. Unfortunately, these quantities predict treatment effect inaccurately because the undirected edges fail to capture biologically plausible causal relations.

A third set of methods utilize directed graphs, where directed edges encode causal relationships. These methods estimate the causal effect of *X*_*i*_ on *Y* via ℙ(*Y* do(*x*_*i*_)), or similarly 𝔼 (*Y* do(*x*_*i*_)), from observational data by conditioning and then marginalizing over an appropriate subset of the variables [20]– [The conditional distribution quantifies the causal effect of *X*_*i*_ on *Y*, but it does not consider the *root* causal effect of *X*_*i*_ on *Y* or the response of the root causal effects to treatment. Accounting for interactions using do-or asymmetric Shapley values fails to rectify the issues [23], [24]. We therefore cannot use these algorithms to discover treatments that mitigate pathogenesis.

We can however summarize the causal effects involved in pathogenesis using the root causal effects, or the total effects of all root causes on a phenotypic response *Y*. Investigators defined the root causal effect of a variable as the predictivity of its error term in a structural equation model [13], [14], [25]. They then proposed to use the Shapley values of [26] – equivalent to the root causal effects in the linear case – in order to identify the root causes of a target vertex. Operationalizing this idea requires invertible SEMs, where we can pinpoint the error term values from the endogenous variables alone. Unfortunately, nature may not obey the bijective relationships needed to recover the error terms exactly. The root causal effects also summarize pathogenic effects but do not quantify their response to treatment.

In this paper, we introduce an algorithm that measures the sample-specific effect of treatment on pathogenesis without relying on the presence of bijective causal relationships. We assume that we can recover a DAG associated with a potentially non-invertible SEM. We then utilize the graph and the endogenous variables alone to recover the root causal effects. Subsequently, we introduce targeted interventions into the DAG and quantify how each intervention changes the total effects from root causes to phenotype *Y* . Clustering of the resultant changes yields groups of patients whose potentially differing pathogeneses respond similarly to treatment.

## IV. Setup

### A. Partially Linear Model

We consider an SEM obeying Equation (1) but enforce a linear SEM in the subset Anc(*Y*) ⊆ ***X*** so that:

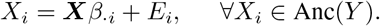

The SEM therefore obeys a partially linear model. The error terms may be Gaussian or discrete, so we do not assume LiNGAM even among Anc(*Y*). The following result holds:

#### Theorem 1.

*The root causal effect of X*_*i*_ *on Y corresponds to the total effect of E*_*i*_ *on Y, or E*_*i*_*θ*_*i*_, *under the partially linear model*.

We delegate proofs to the Appendix, unless explicated in the main text.

### B. Motivating Example

Pathogenesis refers to the development of disease starting from its root causes and ending at its phenotype. We can therefore summarize the pathogenic causal effects using the root causal effects on a phenotypic label *Y* [13]–[15], [25]. Root causal effects may however differ from treatment response. Consider for example the following augmented causal graph:

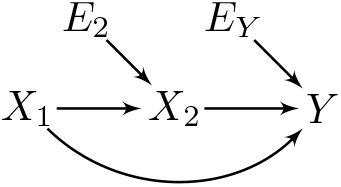

with binary error terms *E*_***1***_ = *X*_***1***_ and *E*_***2***_ whose values are enumerated in Table I (a). We assume that *β* = 1, so that the root causal effects correspond to Table I (b).

**TABLE I:**
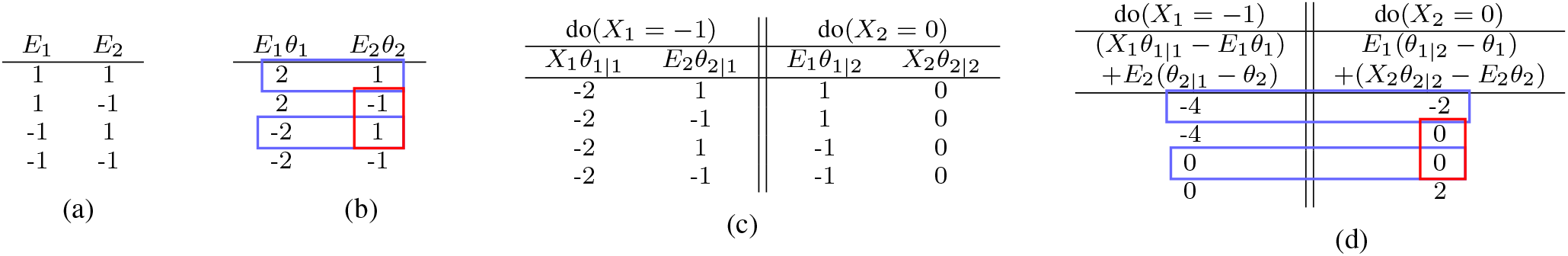
Example of the differences between root causal effects and treatment response.

We next introduce treatments that target specific nodes in ***X***. Let *θ*_*i*|*j*_ denote the total effect of *E*_*i*_ on *Y* after performing do(*X*_*j*_ = *x*_*j*_). If we set do(*X*_***1***_ = − 1), then the root causal effects correspond to the values listed on the left side of Table I (c). Similarly, if we set do(*X*_***2***_ = 0), then we obtain the right side. We list the treatment responses in Table I (d) corresponding to the overall change in root causal effects between Tables I (b) and I (c). We highlight three observations:

1. Different root causal effects may respond similarly to treatment. The red boxes highlight two cases where the root causal effects of *X*_***2***_ are different in Table I (b) but have no treatment response after do(*X*_***2***_ = 0) in Table I (d).
2. Patients may have partially matching root causal effects but entirely different treatment responses. The blue boxes in Table I (b) highlight two cases with the same root causal effects *E*_***2***_*θ*_***2***_. However, the treatment responses for the two cases do not match after either do(*X*_***1***_ = − 1) or do(*X*_***2***_ = 0) in Table I (d).
3. Patients can have non-zero root causal effects but fail to respond to treatment. For example, the second blue box in Table I (b) has non-zero root causal effects but zero treatment responses in Table I (d).

Root causal effects without treatment thus provide insufficient information to predict treatment response. We instead want to quantify the change in root causal effects from before to after treatment.

## V. Strategy

### A. Overview

We now introduce a generalized strategy for identifying the change in root causal effects. We focus on the partially linear model for interpretability. In particular, we consider the *p* error terms that represent potential root causes:

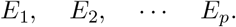

We then weigh each error term by its total effect on a target variable *Y*:

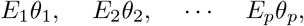

where *θ*_*i*_ = 0 if *X*_*i*_ ∉ Anc(*Y*). We therefore predict the downstream target using the upstream error terms. We will show how to directly recover the above root causal effects even in non-invertible SEMs in the next section.

We next measure the change in the root causal effect of *X*_*i*_ ∈ ***X*** \ *Y* on *Y* after performing do(*X*_*j*_ = *x*_*j*_) with *X*_*j*_ ∈ ***X*** \ *Y*:

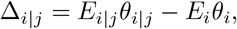

where *E*_*i*|*j*_ = *E*_*i*_ if *X*_*i*_ ≠ *X*_*j*_ and *E*_*i*|*j*_ = *x*_*j*_ otherwise. The quantity 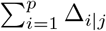 corresponds to the *Treated Root causal Effects* (TRE) quantifying the change in the root causal effects on *Y* after intervening on *X*_*j*_. This process leads to the feature space Π:

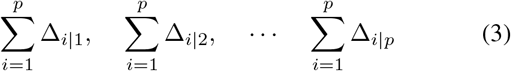

summarizing the TRE for each *X*_*j*_ ∈ ***X*** \ *Y* . If larger values of *Y* correspond to worse symptoms, then we prefer targets associated with negative TREs because they reduce symptoms. We also propose to perform clustering on Π in order to identify theratypes (details in Section V-E).

### B. Endogenous Root Causal Effects

Recovering the root causal effect *E*_*i*_*θ*_*i*_ for each sample requires access to the error term values. We cannot recover the error term values exactly in non-invertible SEMs. We remedy this situation with an alternative approach.

We can write the following using Equation (1):

**Lemma 1**. *We have* ℙ (*Y* |*E*_*i*_, Pa(*X*_*i*_)) = ℙ (*Y* |*X*_*i*_, Pa(*X*_*i*_)). We thus no longer require knowledge of the value of *E*_*i*_ but only the values of the endogenous variables *X*_*i*_ and Pa(*X*_*i*_). Particularizing the above result to conditional expectations yields:

**Corollary 1**. *We have* 𝔼(*Y* |*E*_*i*_, Pa(*X*_*i*_)) = 𝔼(*Y* |*X*_*i*_, Pa(*X*_*i*_)). The above corollary allows us to state:

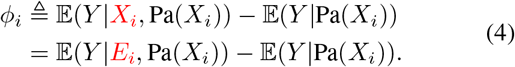

Let ***p*** index Pa(*X*_*i*_) in ***X***. The conditional expectations in the last line correspond to the following under the partially linear model:

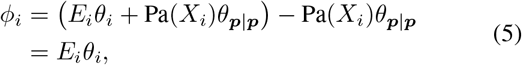

where *θ*_***p***|***p***_ corresponds to the total effect of Pa(*X*_*i*_) on *Y* in 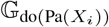. We can therefore compute the root causal effect of *X*_*i*_ on *Y* using the difference 𝔼 (*Y X*_*i*_, Pa(*X*_*i*_)) 𝔼 (*Y* Pa(*X*_*i*_)) relying on endogenous variables alone. We have proved the following main result:

#### Theorem 2.

*The root causal effect of X*_*i*_ *on Y corresponds to ϕ*_*i*_ = 𝔼 (*Y*|*X*_*i*_, Pa(*X*_*i*_)) − 𝔼 (*Y* |Pa(*X*_*i*_)) = *E*_*i*_*θ*_*i*_ *under the partially linear model*.

### C. Treated Root Causal Effects

We now consider the changes to the root causal effects introduced by targeted treatment. Suppose that we set the value of a variable *X*_*j*_ to *x*_*j*_. We quantify the root causal effect of any variable *X*_*i*_ under do(*X*_*j*_ = *x*_*j*_) as follows:

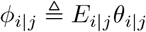

analogous to *ϕ*_*i*_ = *E*_*i*_*θ*_*i*_. The change in the root causal effect after treatment then corresponds to:

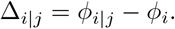

Repeating the above process for every Xi ∈ X \ Y allows us to compute 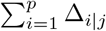 for each *X*_*j*_ ∈ ***X*** \ *Y* and therefore the TREs in Expression (3).

We showed how to compute *ϕ*_*i*_ of each Δ_*i*|*j*_ in the previous section. We can compute *ϕ*_*i*|*j*_ given the coefficient matrix *β* over Anc(*Y*) as follows. Let *θ*_***p***|***p****j*_ correspond to the total effect of Pa(*X*_*i*_) on *Y* in 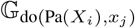. We have:

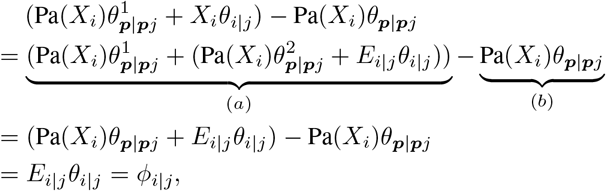

where we have decomposed *θ*_***p***|***p****j*_ into 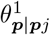 and 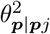 denoting the component of the total effect of Pa(*X*_*i*_) on *Y* in 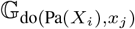 that does not and does pass through *X*_*i*_, respectively. The second equality holds because *X*_*i*_*θ*_*i*|*j*_ is equal to the total effect of Pa(*X*_*i*_) passing through *X*_*i*_ given by 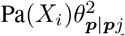 plus the total effect of *E*_*i*_ given by *E*_*i*|*j*_*θ*_*i*|*j*_. We encourage the reader to compare the third line of the above equation to the first line of Equation (5). The terms highlighted by the two underbraces prove the following theorem:

#### Theorem 3.

*ϕ*_*i*|*j*_ *equals (a) the total effect of* Pa(*X*_*i*_) ∪ *X*_*i*_ *on Y in the graph* 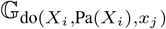 *minus (b) the total effect of* Pa(*X*_*i*_) *on Y in the graph* 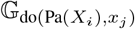 *under the partially linear model*.

We compute the required total effects for any Δ_*i*|*j*_ from the coefficient matrix *β* and therefore recover all of the TREs Π.

### D. Algorithm

Sections V-A through V-C lead to the Root and Treated Root causal Effects (R-TRE) algorithm, which we summarize in Algorithm 1. R-TRE first learns the DAG 𝔾 and corresponding linear coefficient matrix *β* over Anc(*Y*) in Line 1. We assume that we can recover 𝔾 uniquely using any method of choice. In this paper, we use constraint-based search and orient any remaining undirected edges by experimentation or background knowledge [27].

R-TRE then intervenes on each *X*_*j*_ that can block the root causal effect of *X*_*i*_ on *Y* in Line 6; i.e., by intervening on those vertices that are both a descendant of *X*_*i*_ and an ancestor of *Y* (excluding *Y* itself). This allows the algorithm to compute *ϕ*_*i*|*j*_ in Line 6 per Theorem 3. Finally, R-TRE adds in each Δ_*i*|*j*_ in Line 7 in order to recover the TREs Π.

#### Algorithm 1

Root and Treated Root causal Effects (R-TRE)

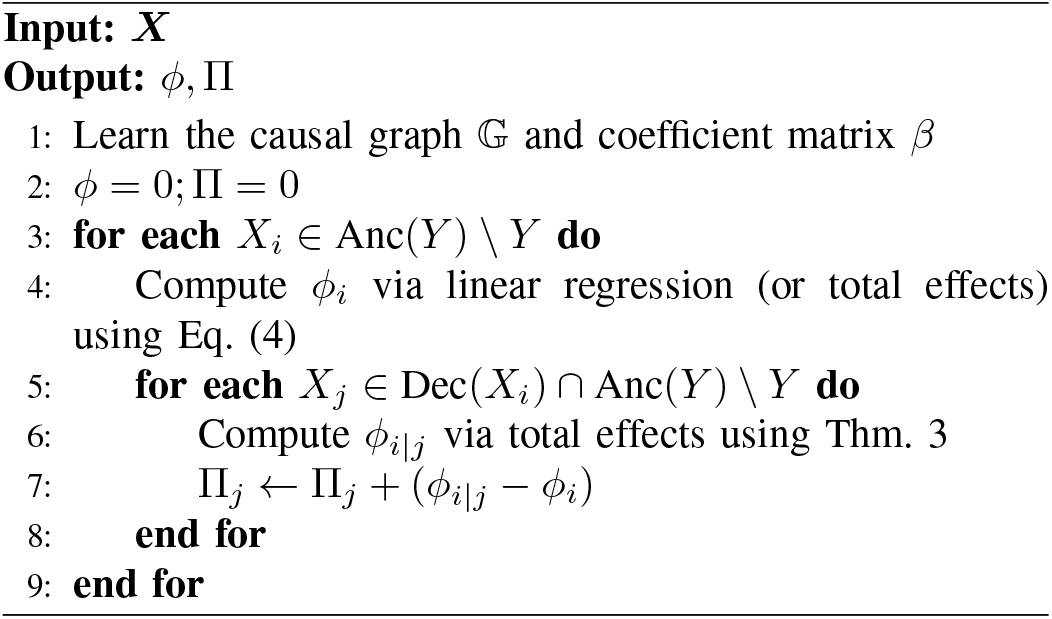

### E. Downstream Clustering

R-TRE outputs the TREs Π corresponding to the changes in the root causal effects after treatment. We can therefore discover theratypes from Π by performing hierarchical clustering on the feature space.

We seek clusters where patients respond similarly to treatment. We therefore perform agglomerative hierarchical clustering on Π using Ward’s method [28]. Ward’s method merges two clusters when the merge leads to a minimum increase in the (weighted) squared Euclidean distance between cluster means. As a result, Ward’s method yields a dendogram where each cluster contains patients who respond similarly to the cluster mean.^1^

The dendogram importantly summarizes nested clusters so that users can identify large enough groups of patients who respond similarly to treatment. Each patient may respond to treatment differently, but categorizations help clinicians quickly comprehend patients by leveraging their past experiences with similar individuals. Recall that the partially linear model of Section IV-A allows discrete error terms that can induce clustering. Clear clusters may not exist in many cases, but we seek to identify them when they do. If a patient does not fall into a cluster, then we resort to a dimensional approach by directly utilizing the recovered TREs [29].

## IV. Other Measures

We emphasize that the TREs recovered by the R-TRE algorithm differ from other measures introduced in the literature. We summarize the discussion in Table II in terms of four criteria – whether the method (1) involves causality, (2) attempts to detect root causes, (3) achieves precision or sample-specificity, or (4) accounts for targeted interventions on the endogenous variables.

**TABLE II:** Comparison against previously proposed measures.

|  | Causality | Root Causes | Precision | Interventions |
| --- | --- | --- | --- | --- |
| MSV |  |  | ✓ |  |
| CSV |  |  | ✓ |  |
| MESV | ✓ | ✓ | ✓ |  |
| ASV | ✓ |  | ✓ |  |
| RCAO | ✓ | ✓ |  |  |
| RCAM | ✓ | ✓ |  |  |
| ICC | ✓ | ✓ | ✓ |  |
| do-Reg | ✓ |  | ✓ | ✓ |
| do-SV | ✓ |  | ✓ | ✓ |
| Reg |  |  | ✓ |  |
| Cor |  |  | ✓ |  |
| TRE | ✓ | ✓ | ✓ | ✓ |

### Conditional and Marginal Shapley Values (CSV, MSV)

[26], [30] These Shapley values quantify feature importance according to a trained model. The conditional and marginal Shapley values marginalize over the inputs of the model using conditional or marginal expectations, respectively. Authors have argued that marginal Shapley values better represent causal relations between *algorithm* input-output pairs [30]. Marginal Shapley values however do not represent the causality in *nature* required for TREs and disease subtyping.

### Marginal Error term Shapley Values (MESV)

[13]–[15], [25] These features correspond to marginal Shapley values on the error terms. Marginal error term Shapley values are equivalent to the root causal effects under a linear SEM provided that we can recover the error term values uniquely under invertibility. R-TRE recovers root causal effects even in the non-invertible scenario.

### Asymmetric Shapley Values (ASV)

[23] Asymmetric Shapley values are conditional Shapley values that take into account natural causality by marginalizing over subsets of the ancestors of each variable. Asymmetric Shapley values therefore incorporate variable ordering. In contrast, R-TRE *always* conditions on *all* of the parents in order to explicitly recover root causal effects and TREs.

### Root Causal Analysis of Outliers (RCAO) or Marginals (RCAM)

[31], [32] These values quantify the contribution of the error terms on an outlier score or a marginal distribution. They therefore require an invertible SEM, forego sample-specificity and do not quantify root causal effects under interventions.

### Intrinsic Causal Contribution (ICC)

[33] ICC quantifies the reduction in uncertainty of a variable after knowing the value of an error term. This method therefore again assumes that we can recover the error term values uniquely from an invertible SEM.

### do-Regression (do-Reg) and do-Shapley Values (do-SV)

[24], [34] Do-regression computes 𝔼 (*Y* |do(*X*_*i*_)) for each *X*_*i*_ ∈ ***X* \** *Y* whereas do-Shapley values perform the do-operation across all subsets of ***X* \** *Y* . These algorithms perform interventions but do not specifically track for changes in root causal effects.

### Standard Regression (Reg) and Correlation (Cor)

We include standard multivariate linear regression and correlation (or univariate linear regression) as sanity checks. We weigh each feature by its regression coefficient.

We conclude that only TREs account for the change in root causal effects after performing targeted interventions, and R-TRE does not require an invertible SEM.

## VII. Experiments

We now evaluate the accuracy of R-TRE and compare it against other algorithms recovering the measures of Section VI.

### A. Data Generation

We first generated a linear SEM obeying Equation (2) with *p* = 10 variables. We created the coefficient matrix *β* by sampling from a Bernoulli(2*/*(*p*− 1)) distribution in the upper triangular portion of the matrix with an expected neighborhood size of 2. We then randomly permuted the variable ordering. We introduced weights *β* by uniformly sampling from [−1, −0.25] ∪ [0.25, 1]. We chose the distributions of the error terms uniformly at random from the following set: a uniform distribution between -1 and 1, a t-distribution with three degrees of freedom, or a discrete uniform distribution with 2 or 3 values. We centered all error terms. We then binarized a random subset of the variables with at most one child to ensure a non-invertible SEM. We chose *Y* randomly from the set of random variables with at least one parent; *Y* need not be a terminal vertex. We repeated the above procedure 250 times for sample sizes 1000, 3000 and 9000. We therefore generated a total of 250 × 3 = 750 unique datasets.

#### Reproducibility

All code needed to reproduce the experimental results is available at https://github.com/ericstrobl/RTRE.

### B. Comparators & Metric

We compare the output of R-TRE against algorithms recovering the eleven other measures listed in Table II. We learned the DAG for all algorithms using the PC algorithm equipped with the linear correlation test and an alpha threshold of 0.01 [27].

We evaluated the accuracy in recovering the root causal effects and the TREs using the root mean squared error (RMSE) to the ground truth:

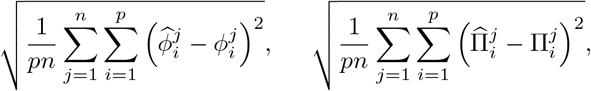

where the superscripts index the *n* samples. We also evaluated the accuracy of the clustered TREs using the RMSE:

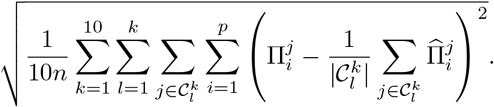

The set 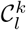 contains the sample indices of the *l*^th^ cluster among a total of *k* clusters. We vary *k* from 1 to 10. The above metric quantifies the distance from the true TREs to their estimated cluster means, i.e., the estimated treatment response by group.

We collectively call these cluster means the *Clustered Treated Root causal Effects* (CTRE). We plot mean sum of squares (squared RMSE) for each *k* when varying the value of *k* in order to conform with tradition [28]. Lower values of the above three metrics denote better performance.

### C. Results

We summarize the accuracy results in Table III. Bolded values denote the best performance according to a paired t-test significant at the Bonferonni corrected threshold of 0.05/12, since we compared a total of 12 algorithms. We place timing results in Table IV in the Appendix; R-TRE always completed within 10 milliseconds on average.

**TABLE III:**
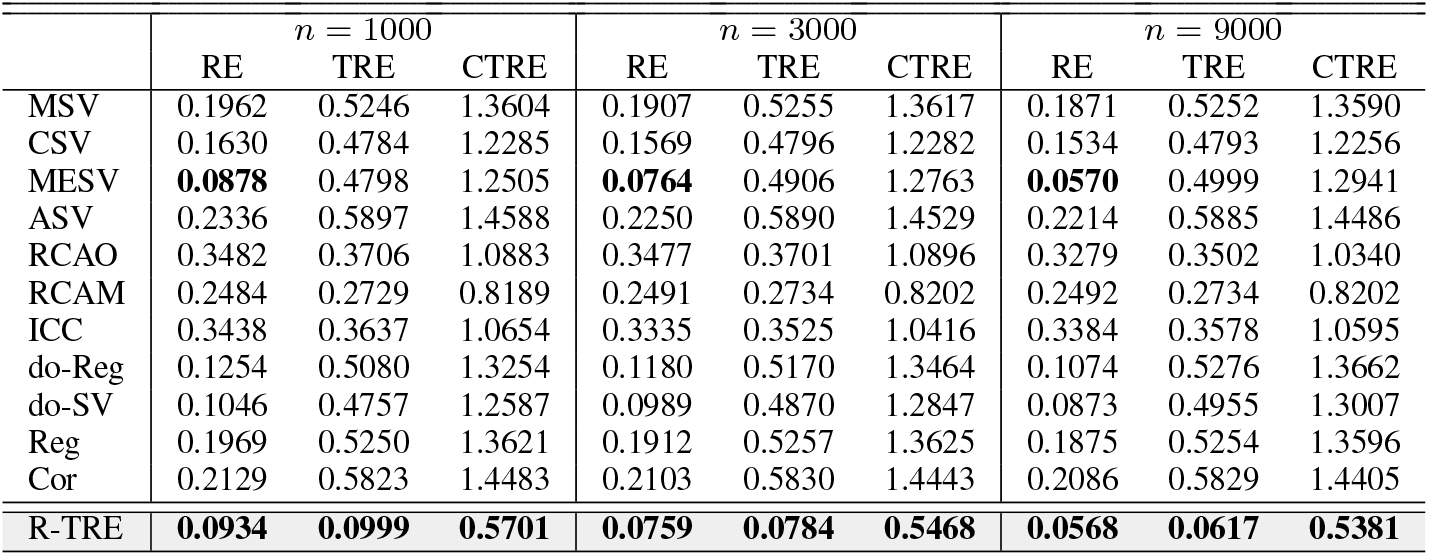
Mean RMSE results to the ground truth RE, TRE and CTRE values. Bolded values highlight the best performances. R-TRE achieves the lowest RMSE in all three cases across all sample sizes.

| | $n = 1000$ | | | $n = 3000$ | | | $n = 9000$ | | |
| --- | --- | --- | --- | --- | --- | --- | --- | --- | --- |
|  | RE | TRE | CTRE | RE | TRE | CTRE | RE | TRE | CTRE |
| MSV | 0.1962 | 0.5246 | 1.3604 | 0.1907 | 0.5255 | 1.3617 | 0.1871 | 0.5252 | 1.3590 |
| CSV | 0.1630 | 0.4784 | 1.2285 | 0.1569 | 0.4796 | 1.2282 | 0.1534 | 0.4793 | 1.2256 |
| MESV | **0.0878** | 0.4798 | 1.2505 | **0.0764** | 0.4906 | 1.2763 | **0.0570** | 0.4999 | 1.2941 |
| ASV | 0.2336 | 0.5897 | 1.4588 | 0.2250 | 0.5890 | 1.4529 | 0.2214 | 0.5885 | 1.4486 |
| RCAO | 0.3482 | 0.3706 | 1.0883 | 0.3477 | 0.3701 | 1.0896 | 0.3279 | 0.3502 | 1.0340 |
| RCAM | 0.2484 | 0.2729 | 0.8189 | 0.2491 | 0.2734 | 0.8202 | 0.2492 | 0.2734 | 0.8202 |
| ICC | 0.3438 | 0.3637 | 1.0654 | 0.3335 | 0.3525 | 1.0416 | 0.3384 | 0.3578 | 1.0595 |
| do-Reg | 0.1254 | 0.5080 | 1.3254 | 0.1180 | 0.5170 | 1.3464 | 0.1074 | 0.5276 | 1.3662 |
| do-SV | 0.1046 | 0.4757 | 1.2587 | 0.0989 | 0.4870 | 1.2847 | 0.0873 | 0.4955 | 1.3007 |
| Reg | 0.1969 | 0.5250 | 1.3621 | 0.1912 | 0.5257 | 1.3625 | 0.1875 | 0.5254 | 1.3596 |
| Cor | 0.2129 | 0.5823 | 1.4483 | 0.2103 | 0.5830 | 1.4443 | 0.2086 | 0.5829 | 1.4405 |
| R-TRE | **0.0934** | **0.0999** | **0.5701** | **0.0759** | **0.0784** | **0.5468** | **0.0568** | **0.0617** | **0.5381** |

**TABLE IV:**
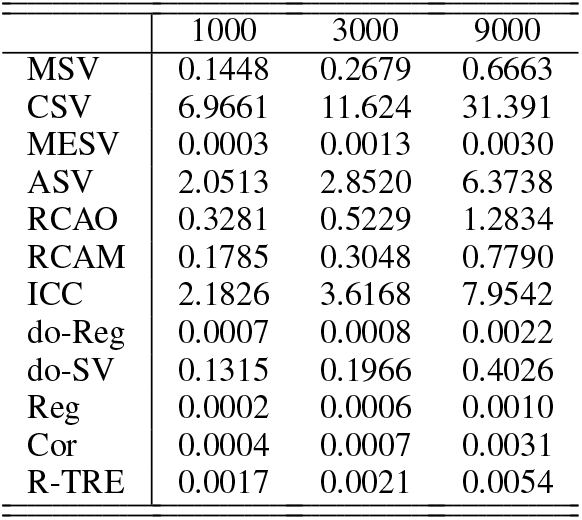
Timing results for the synthetic data in seconds. Columns correspond to different sample sizes.

|  | 1000 | 3000 | 9000 |
| --- | --- | --- | --- |
| MSV | 0.1448 | 0.2679 | 0.6663 |
| CSV | 6.9661 | 11.624 | 31.391 |
| MESV | 0.0003 | 0.0013 | 0.0030 |
| ASV | 2.0513 | 2.8520 | 6.3738 |
| RCAO | 0.3281 | 0.5229 | 1.2834 |
| RCAM | 0.1785 | 0.3048 | 0.7790 |
| ICC | 2.1826 | 3.6168 | 7.9542 |
| do-Reg | 0.0007 | 0.0008 | 0.0022 |
| do-SV | 0.1315 | 0.1966 | 0.4026 |
| Reg | 0.0002 | 0.0006 | 0.0010 |
| Cor | 0.0004 | 0.0007 | 0.0031 |
| R-TRE | 0.0017 | 0.0021 | 0.0054 |

#### Root Causal Effects

Both R-TRE and MESV achieved the lowest RMSE in discovering the root causal effects (REs). MESV however requires access to the error terms, whereas R-TRE discovers the root causal effects using endogenous variables alone. The do-Shapley values came in third place and struggled to improve beyond the lowest tested sample size. We conclude that R-TRE accurately discovers sample-specific REs.

#### Treated Root Causal Effects

R-TRE outperformed all other algorithms by a large margin in estimating TREs; RCAM came in second place but with greater than 2.5 times the error. We conclude that R-TRE also accurately discovers sample-specific TREs.

#### Clustered Treated Root Causal Effects

Clustering results in loss of information due to the groupings. R-TRE however still outperformed all algorithms after clustering the estimated TREs from one to ten groups. We plot the accuracy results of the top three algorithms for up to 10 total clusters in Figure 1 for *n* = 9000; RCAM had a flat sum of squares line because its output is not sample-specific. Only the performance of R-TRE continued to improve with increasing numbers of clusters. We conclude that R-TRE accurately identifies samples with similar TREs regardless of total cluster number.

**Fig. 1:**
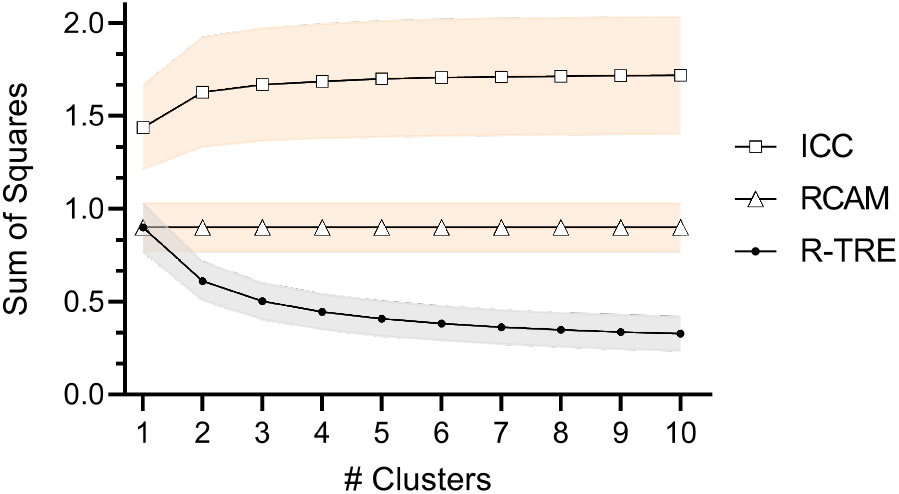
Mean sum of squares across different cluster sizes for first, second and third place algorithms at *n* = 9000. Bands denote 95% confidence intervals. R-TRE continues to improve with increasing numbers of clusters, but the other two algorithms do not.

In summary, R-TRE outputs accurate REs, TREs and CTREs for target discovery. The algorithm outperforms all 11 other algorithms.

### D. Applications

We demonstrate the generality of the approach by applying R-TRE and the other algorithms to a clinical dataset, a single-cell RNA sequencing (scRNA-seq) dataset and a flow cytometry dataset. We report the main results here but refer the reader to the Appendix for full tables of accuracy and timing results. We equipped the PC algorithm with a fast non-parametric conditional independence test for all real datasets [13], [35].

#### Type II Diabetes

We sought to identify TREs and theratypes in Type II diabetes using a real clinical dataset [36].^2^ The dataset contains 7 variables related to the metabolic system in 768 Pima Indians; we excluded insulin due to a possible cycle involving insulin and glucose. Type II diabetes is well-studied, so we expect R-TRE to only identify one cluster of patients using clinical variables [37]. Note that multiple theratypes of type II diabetes likely exist, but they are not detectable when intervening on the routine clinical variables present in this dataset [38].

The dataset comes with a known ground-truth graph, which we used to fit the parameters of a linear SEM to obtain the ground truth [15]. We then ran all of the algorithms on 250 bootstrapped draws of the dataset. The algorithms only had access to the graph estimated from bootstrapped samples using PC. We did not intervene on age and pedigree for R-TRE, do-reg and do-SV, since we cannot intervene on these variables in practice.

We summarize results in Figures 2 (a) and 2 (b). R-TRE achieved the lowest mean RMSE to the ground truth TREs as compared to the 11 other algorithms (Figure 2 (a)). We plot the clustering results of the top two algorithms including R-TRE and RCAM in Figure 2 (b); the sum of squares for all other algorithms increased with increasing number of clusters implying worse performance. The clustering results of R-TRE in Figure 2 (b) show a sharp drop in the sum of squares after one cluster and a subsequent leveling off. R-TRE therefore only identified approximately one theratype in this dataset as expected. We conclude that R-TRE identified the correct number of clusters and estimated treatment effect most accurately in the Pima Indians dataset.

**Fig. 2:**
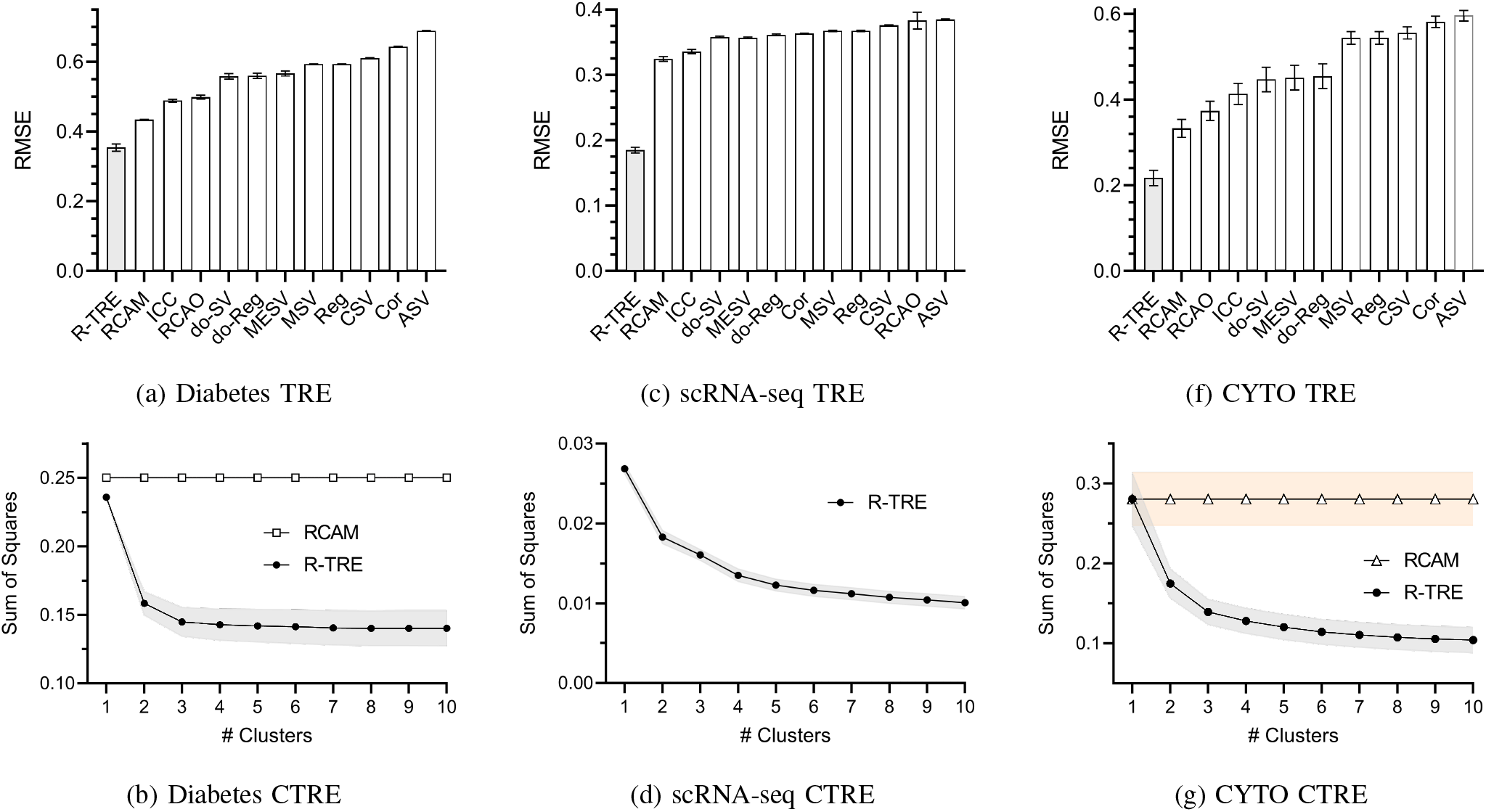
TRE and CTRE accuracy results for the real datasets. Error bars and bands again denote 95% confidence intervals.

#### Single-Cell RNA Sequencing in Disease

We next sought to increase the difficulty by utilizing an scRNA-seq dataset of lung adenocarcinoma [39].^3^ R-TRE is sample-specific in general, so we can use the algorithm to identify TREs of individual cells rather than just patients. The scRNA-seq dataset contains 17,502 single cells derived from cancerous and normal adjacent tissue in three individuals. We thus expect to detect *at least* three clusters of cells due to the heterogeneity of the cell population. We focused on variables involved in the mitogen-activated protein kinase (MAPK) pathway because of its importance in lung cancer pathogenesis. The variables include lung adenocarcinoma status as well as expression levels of GRB2, HRAS, ARAF, CCND1. We also included KRAS and TP53. We excluded EGFR since it had nearly all zero counts in the dataset. We extracted the ground truth causal graph from the KEGG pathway of non-small cell lung cancer (HSA05223) [40], [41].

We set lung adenocarconima status as the target. We report the results in the same format as with the previous example over 100 bootstrapped draws in Figures 2 (c) and 2 (d). R-TRE again estimated the TREs to the highest accuracy. Moreover, clustering results showed a gradual decline in the sum of squares rather than a leveling off. R-TRE identified at least 4 sub-populations of cells using the elbow method. We do not plot any other algorithms in Figure 2 (d) because they all performed much worse than R-TRE. We conclude that R-TRE achieved the highest accuracy and expected clustering results in this dataset.

#### Cell Signaling

The above two datasets use a diagnosis as a discrete terminal vertex. In this example, we demonstrate that R-TRE works well even if the target is continuous and non-terminal. We used the CYTO dataset which contains measurements of 11 phosphoproteins and phospholipids from 7466 primary human immune system cells across 9 experimental conditions [42], [43].^4^ We standardized the data by mean and standard deviation in each experimental condition. The dataset again comes with a ground truth causal graph to fit a linear SEM. We chose the target uniformly at random for vertices that contain at least one parent. We do not expect to see a clear number of clusters in this case, since we vary the target. The inability to find well-defined clusters informs the user to examine the sample TRE values rather than adopt a categorical approach.

We summarize the results over 250 bootstrapped repetitions in Figures 2 (f) and 2 (g). R-TRE again achieved the lowest RMSE to the TREs by a large margin. Furthermore, clustering revealed a smooth decay in the sum of squares, suggesting that either many small or no meaningful groups exist in the data (Figure 2 (g)). We conclude that R-TRE works well across a variety of response variables, even if the cells fail to cluster into a few groups.

In summary, real data results indicate that R-TRE performs well across a variety of scenarios. The algorithm estimates the root causal effects and TREs accurately; it also identifies the theratypes we expect to see across three different dataset types.

## VIII. Conclusion

We summarized the causal effects involved in pathogenesis using root causal effects. We then quantified the response of the pathogenic mechanisms to treatment using TREs. We discovered theratypes by clustering samples with similar TREs. We finally automated these three ideas under a partially linear model with the R-TRE algorithm that simultaneously recovers root causal effects, TREs and CTREs. The algorithm importantly recovers the above quantities even in non-invertible SEMs, where we cannot recover the error term values exactly. Experimental results revealed substantially increased accuracy relative to existing algorithms. Future work will focus on extending R-TRE to the non-linear setting, accommodating confounding as well as incorporating high throughput experimentation in large scRNA-seq datasets.

## Data Availability

All data produced in the present work are contained in the manuscript

https://github.com/ericstrobl/RTRE

## APPENDIX

*Proofs*

### Theorem 1.

*Proof*. We consider the Shapley value formulation introduced in [13]–[15], where:

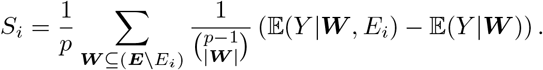

We have *Y* = ***E***_Anc(*Y)*_*θ*_Anc(*Y)*_+ ***E***_***N***_ *θ*_***N***_ under the partially linear model, where *θ* corresponds to the total effects of the error terms on *Y*, ***N*** = ***X*** *\* Anc(*Y*) and *θ*_***N***_ = 0. We now invoke Corollary 1 of [26] to conclude that the root causal effect of *X*_*i*_ corresponds to *S*_*i*_ = *E*_*i*_*θ*_*i*_. □

### Lemma 1.

*We have* ℙ(*Y* |*E*_*i*_, Pa(*X*_*i*_)) = ℙ(*Y* |*X*_*i*_, Pa(*X*_*i*_)).

*Proof*. We can write the following sequence of equalities:

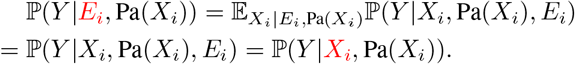

The second equality follows because *X*_*i*_ is a constant given *E*_*i*_ and Pa(*X*_*i*_). We justify the last equality on a case-by-case basis:

1. If *X*_*i*_ is an ancestor of *Y*, then the last equality holds because *E*_*i*_ and *Y* are conditionally independent given *X*_*i*_ ∪ Pa(*X*_*i*_) by the global Markov property.
2. If *X*_*i*_ is not an ancestor of *Y*, then we have two subcases. If *Y ∈* Pa(*X*_*i*_), then *E*_*i*_ and *Y* are conditionally independent given *X*_*i*_ ∪ Pa(*X*_*i*_) because *Y* is a constant given Pa(*X*_*i*_). If *Y ∉* Pa(*X*_*i*_), then again *E*_*i*_ and *Y* are conditionally independent given *X*_*i*_ ∪ Pa(*X*_*i*_) by the global Markov property because *E*_*i*_ and *Y* are d-separated given *X*_*i*_ ∪ Pa(*X*_*i*_).

We have considered all cases, whence the conclusion follows.

*Other Experimental Results*

**TABLE V:**
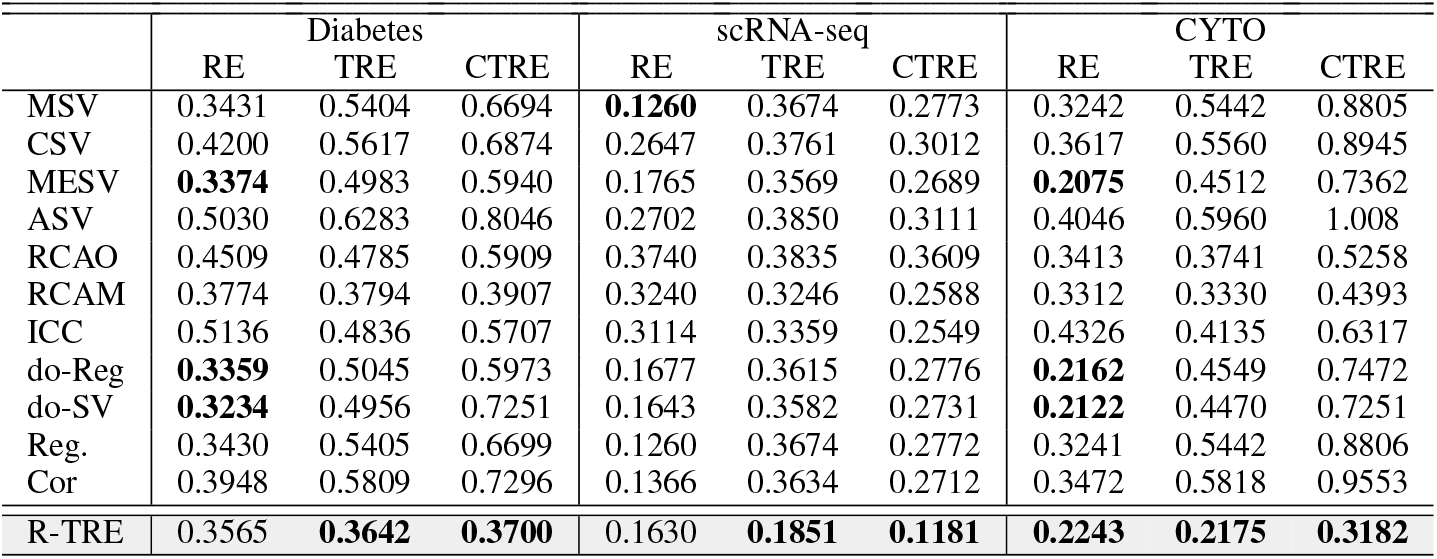
Accuracy results for the real data.

|  | Diabetes |  |  | scRNA-seq |  |  | CYTO |  |  |
| --- | --- | --- | --- | --- | --- | --- | --- | --- | --- |
|  | RE | TRE | CTRE | RE | TRE | CTRE | RE | TRE | CTRE |
| MSV | 0.3431 | 0.5404 | 0.6694 | **0.1260** | 0.3674 | 0.2773 | 0.3242 | 0.5442 | 0.8805 |
| CSV | 0.4200 | 0.5617 | 0.6874 | 0.2647 | 0.3761 | 0.3012 | 0.3617 | 0.5560 | 0.8945 |
| MESV | **0.3374** | 0.4983 | 0.5940 | 0.1765 | 0.3569 | 0.2689 | **0.2075** | 0.4512 | 0.7362 |
| ASV | 0.5030 | 0.6283 | 0.8046 | 0.2702 | 0.3850 | 0.3111 | 0.4046 | 0.5960 | 1.008 |
| RCAO | 0.4509 | 0.4785 | 0.5909 | 0.3740 | 0.3835 | 0.3609 | 0.3413 | 0.3741 | 0.5258 |
| RCAM | 0.3774 | 0.3794 | 0.3907 | 0.3240 | 0.3246 | 0.2588 | 0.3312 | 0.3330 | 0.4393 |
| ICC | 0.5136 | 0.4836 | 0.5707 | 0.3114 | 0.3359 | 0.2549 | 0.4326 | 0.4135 | 0.6317 |
| do-Reg | **0.3359** | 0.5045 | 0.5973 | 0.1677 | 0.3615 | 0.2776 | **0.2162** | 0.4549 | 0.7472 |
| do-SV | **0.3234** | 0.4956 | 0.7251 | 0.1643 | 0.3582 | 0.2731 | **0.2122** | 0.4470 | 0.7251 |
| Reg. | 0.3430 | 0.5405 | 0.6699 | 0.1260 | 0.3674 | 0.2772 | 0.3241 | 0.5442 | 0.8806 |
| Cor | 0.3948 | 0.5809 | 0.7296 | 0.1366 | 0.3634 | 0.2712 | 0.3472 | 0.5818 | 0.9553 |
| R-TRE | 0.3565 | **0.3642** | **0.3700** | 0.1630 | **0.1851** | **0.1181** | **0.2243** | **0.2175** | **0.3182** |

**TABLE VI:** Timing results for the real data in seconds.

|  | Diabetes | scRNA-seq | CYTO |
| --- | --- | --- | --- |
| MSV | 0.0946 | 0.5997 | 0.6707 |
| CSV | 5.3320 | 29.653 | 45.676 |
| MESV | 0.0002 | 0.0043 | 0.0022 |
| ASV | 1.8635 | 7.2489 | 7.1502 |
| RCAO | 0.4762 | 5.6888 | 0.9319 |
| RCAM | 0.2501 | 2.6468 | 0.3252 |
| ICC | 2.5110 | 41.712 | 7.2634 |
| do-Reg | 0.0006 | 0.0048 | 0.0022 |
| do-SV | 0.0389 | 0.3003 | 0.4386 |
| Reg | 0.0002 | 0.0024 | 0.0013 |
| Cor | 0.0004 | 0.0038 | 0.0025 |
| R-TRE | 0.0018 | 0.0201 | 0.0038 |

## Footnotes

1 We focus on interpretability with a dendogram, but users may employ alternative clustering methods depending on the needs of their particular application. The clusters must group patients who respond similarly to treatment.

2 https://www.kaggle.com/datasets/uciml/pima-indians-diabetes-database

3 https://www.ncbi.nlm.nih.gov/geo/query/acc.cgi?acc=GSE123904

4 https://arxiv.org/src/1805.03108v1/anc/data.txt

## References

G. W. Imbens and D. B. Rubin, Causal Inference in Statistics, Social, and Biomedical Sciences. Cambridge University Press, 2015.

J. Pearl, Causality. Cambridge: Cambridge University Press, 2009.

A. Bhangu, K. Søreide, S. Di Saverio, J. H. Assarsson, and F. T. Drake, “Acute appendicitis: Modern understanding of pathogenesis, diagnosis, and management,” The Lancet, vol. 386, no. 10000, pp. 1278–1287, 2015.

K. B. Wray, Resisting Scientific Realism. Cambridge University Press, 2018.

K. Jaspers, General Psychopathology. Johns Hopkins University Press, 1997, vol. 2.

I. Kant, J. M. D. Meiklejohn, T. K. Abbott, and J. C. Meredith, Critique of Pure Reason. JM Dent London, 1934.

I. Agache, C. A. Akdis, et al., “Precision medicine and phenotypes, endotypes, genotypes, regiotypes, and theratypes of allergic diseases,” The Journal of Clinical Investigation, vol. 129, no. 4, pp. 1493–1503, 2019.

A. D. Sheftel, A. B. Mason, and P. Ponka, “The long history of iron in the universe and in health and disease,” Biochimica et Biophysica Acta (BBA)-General Subjects, vol. 1820, no. 3, pp. 161–187, 2012.

H. F. Bunn, “Vitamin b12 and pernicious anemia - the dawn of molecular medicine,” New England Journal of Medicine, vol. 370, no. 8, pp. 773–776, 2014.

K. Zhang, Z. Wang, J. Zhang, and B. Schölkopf, “On estimation of functional causal models: General results and application to the post-nonlinear causal model,” ACM Transactions on Intelligent Systems and Technology (TIST), vol. 7, no. 2, pp. 1–22, 2015.

S. L. Lauritzen, A. P. Dawid, B. N. Larsen, and H.-G. Leimer, “Independence properties of directed markov fields,” Networks, vol. 20, no. 5, pp. 491–505, 1990.

S. Shimizu, P. O. Hoyer, A. Hyvärinen, A. Kerminen, and M. Jordan, “A linear non-gaussian acyclic model for causal discovery.,” Journal of Machine Learning Research, vol. 7, no. 10, 2006.

E. V. Strobl, “Counterfactual formulation of patient-specific root causes of disease,” arXiv preprint 2305.17574, 2023.

E. V. Strobl and T. A. Lasko, “Identifying patient-specific root causes of disease,” in Proceedings of the 13th ACM International Conference on Bioinformatics, Computational Biology and Health Informatics, ser. BCB’ 22, Northbrook, Illinois: Association for Computing Machinery, 2022, ISBN: 9781450393867.

E. V. Strobl and T. A. Lasko, “Sample-specific root causal inference with latent variables,” Causal Learning and Reasoning, 2023.

R. Macarron, M. N. Banks, D. Bojanic, et al., “Impact of highthroughput screening in biomedical research,” Nature Reviews Drug Discovery, vol. 10, no. 3, pp. 188–195, 2011.

E. E. Schadt, S. H. Friend, and D. A. Shaywitz, “A network view of disease and compound screening,” Nature Reviews Drug Discovery, vol. 8, no. 4, pp. 286–295, 2009.

A.-L. Barabási, N. Gulbahce, and J. Loscalzo, “Network medicine: A network-based approach to human disease,” Nature Reviews Genetics, vol. 12, no. 1, pp. 56–68, 2011.

E. Guney, J. Menche, M. Vidal, and A.-L. Barábasi, “Network-based in silico drug efficacy screening,” Nature Communications, vol. 7, no. 1, p. 10 331, 2016.

M. H. Maathuis, M. Kalisch, and P. Bühlmann, “Estimating highdimensional intervention effects from observational data,” The Annals of Statistics, vol. 37, no. 6A, pp. 3133–3164, 2009. DOI: 10.1214/09-AOS685. [Online]. Available: https://doi.org/10.1214/09-AOS685.

D. Malinsky and P. Spirtes, “Estimating causal effects with ancestral graph markov models,” in Conference on Probabilistic Graphical Models, PMLR, 2016, pp. 299–309.

T.-Z. Wang, T. Qin, and Z.-H. Zhou, “Sound and complete causal identification with latent variables given local background knowledge,” Advances in Neural Information Processing Systems, vol. 35, pp. 10 325–10338, 2022.

C. Frye, C. Rowat, and I. Feige, “Asymmetric shapley values: Incorporating causal knowledge into model-agnostic explainability,” Advances in Neural Information Processing Systems, vol. 33, pp. 1229–1239, 2020.

Y. Jung, S. Kasiviswanathan, J. Tian, D. Janzing, P. Blöbaum, and E. Bareinboim, “On measuring causal contributions via do-interventions,” in International Conference on Machine Learning, PMLR, 2022, pp. 10 476–10 501.

E. V. Strobl and T. A. Lasko, “Identifying patient-specific root causes with the heteroscedastic noise model,” Journal of Computational Science, vol. 72, 2023.

S. M. Lundberg and S.-I. Lee, “A unified approach to interpreting model predictions,” in Proceedings of the 31st International Conference on Neural Information Processing Systems, 2017, pp. 4768–4777.

P. Spirtes, C. Glymour, and R. Scheines, Causation, Prediction, and Search, 2nd. MIT press, 2000.

J. H. Ward Jr, “Hierarchical grouping to optimize an objective function,” Journal of the American Statistical Association, vol. 58, no. 301, pp. 236–244, 1963.

V. Peralta and M. J. Cuesta, “A dimensional and categorical architecture for the classification of psychotic disorders,” World Psychiatry, vol. 6, no. 2, p. 100, 2007.

D. Janzing, L. Minorics, and P. Blöbaum, “Feature relevance quantification in explainable ai: A causal problem,” in International Conference on Artificial Intelligence and Statistics, PMLR, 2020, pp. 2907–2916.

K. Budhathoki, D. Janzing, P. Bloebaum, and H. Ng, “Why did the distribution change?” In International Conference on Artificial Intelligence and Statistics, PMLR, 2021, pp. 1666–1674.

K. Budhathoki, L. Minorics, P. Blöbaum, and D. Janzing, “Causal structure-based root cause analysis of outliers,” in International Conference on Machine Learning, PMLR, 2022, pp. 2357–2369.

D. Janzing, P. Blöbaum, L. Minorics, P. Faller, and A. Mastakouri, “Quantifying intrinsic causal contributions via structure preserving interventions,” arXiv e-prints, 2020.

T. Heskes, E. Sijben, I. G. Bucur, and T. Claassen, “Causal shapley values: Exploiting causal knowledge to explain individual predictions of complex models,” Advances in Neural Information Processing Systems, vol. 33, pp. 4778–4789, 2020.

E. V. Strobl, K. Zhang, and S. Visweswaran, “Approximate kernel-based conditional independence tests for fast non-parametric causal discovery,” Journal of Causal Inference, vol. 7, no. 1, 2019.

J. W. Smith, J. E. Everhart, W. Dickson, W. C. Knowler, and R. S. Johannes, “Using the adap learning algorithm to forecast the onset of diabetes mellitus,” in Proceedings of the Annual Symposium on Computer Application in Medical Care, American Medical Informatics Association, 1988, p. 261.

M. J. Davies, V. R. Aroda, B. S. Collins, et al., “Management of hyperglycemia in type 2 diabetes, 2022. a consensus report by the american diabetes association (ada) and the european association for the study of diabetes (easd),” Diabetes Care, vol. 45, no. 11, pp. 2753–2786, 2022.

S. Schrader, A. Perfilyev, E. Ahlqvist, et al., “Novel subgroups of type 2 diabetes display different epigenetic patterns that associate with future diabetic complications,” Diabetes Care, vol. 45, no. 7, pp. 1621–1630, 2022.

A. M. Laughney, J. Hu, N. R. Campbell, et al., “Regenerative lineages and immune-mediated pruning in lung cancer metastasis,” Nature Medicine, vol. 26, no. 2, pp. 259–269, 2020.

M. Kanehisa and S. Goto, “Kegg: Kyoto encyclopedia of genes and genomes,” Nucleic Acids Research, vol. 28, no. 1, pp. 27–30, 2000.

E. V. Strobl and T. A. Lasko, “Root causal inference from single cell rna sequencing with the negative binomial,” in Proceedings of the 14th ACM International Conference on Bioinformatics, Computational Biology and Health Informatics, ser. BCB’ 23, Northbrook, Illinois: Association for Computing Machinery, 2023.

K. Sachs, O. Perez, D. Pe’er, D. A. Lauffenburger, and G. P. Nolan, “Causal protein-signaling networks derived from multiparameter single-cell data,” Science, vol. 308, no. 5721, pp. 523–529, 2005.

J. Ramsey and B. Andrews, “Fask with interventional knowledge recovers edges from the sachs model,” arXiv preprint 1805.03108, 2018.

